# Identifying Family Relationships from Electronic Health Records: A Machine Learning Approach

**DOI:** 10.64898/2026.09.18.26363428

**Authors:** Abhinav Pundir, Andrew Hill, Michael G. Kahn, Bethany M. Kwan, Daniel M. Lindberg, Shaun J. Grannis, Titus K. Schleyer, Lisa M. Schilling, Sarah V. Kautz, Toan C. Ong

**Affiliations:** Colorado School of Public Health, University of Colorado Anschutz, Aurora, CO 80045, United States; Department of Emergency Medicine, Kempe Center for the Prevention & Treatment of Child Abuse & Neglect, University of Colorado Anschutz, Aurora, CO 80045, United States; Department of Family Medicine, Indiana University School of Medicine, Indianapolis, Indiana, United States; Regenstrief Institute, Indianapolis, IN, United States; Division of General Internal Medicine, University of Colorado Anschutz, Aurora, CO 80045, United States; Colorado Clinical and Translational Sciences Institute, University of Colorado Anschutz, Aurora, CO 80045, United States; Department of Biomedical Informatics, University of Colorado Anschutz, Aurora, CO 80045, United States

**Keywords:** family linkage, electronic health records, machine learning, Random Forest, record linkage, patient-centered outcomes research, health informatics

## Abstract

**Objective:** To develop and evaluate machine learning methods for automated identification of family relationships from electronic health records (EHRs). We employed Random Forest classifiers to identify five relationship types: Mother-Child, Father-Child, Sibling-Sibling, Twin-Twin, and Partner-Partner, using a four-stage iterative refinement process to improve precision for patient-centered outcomes research (PCOR) and healthcare applications.

**Materials and Methods:** We used two large-scale Indiana datasets: the Indiana Network for Patient Care (INPC), comprising approximately 15 million unique individuals across approximately 45 million medical records, and the Indiana Natality dataset (birth certificates, 1970-2024), which served as the gold standard. Positive cases were derived by linking verified relationships from Natality records to INPC using a shared Global Identifier. Negative cases were drawn from a three-tier blocking strategy combined with sliding window restriction and similarity scoring, which together reduced the comparison space from over 10¹⁴ potential pairs to approximately 121 million candidates. Five Random Forest classifiers underwent four-stage iterative refinement: initial training, feature removal for multicollinearity, feature engineering to address systematic errors, and training-data refinement restricted to relationship pairs with at least one shared contact feature.

**Results:** The models achieved precision of 0.92 to 1.00, recall of 0.97 to 1.00, and F1 scores of 0.94 to 1.00 across all relationship types (Mother-Child 0.97, Father-Child 0.98, Sibling-Sibling 0.98, Twin-Twin 1.00, Partner-Partner 0.94). Between 15% and 78% of Natality-verified relationships lacked any shared contact information in INPC and were structurally undetectable by record linkage; restricting training and evaluation to linkable pairs improved precision for four of the five models, and training-data refinement improved F1-scores by a further 0.02-0.04 for the Mother-Child, Father-Child, and Sibling models. High-confidence predictions (probability ≥ 0.9) captured 77-99% of true positives. Age difference was the primary predictor for parent-child and twin relationships, while phone number similarity was most important for siblings.

**Discussion:** The framework identifies family relationships from standard demographic fields available in most EHR systems. The iterative refinement approach shows how systematic error analysis can guide methodological improvements. Training-data refinement addresses a constraint in EHR-based family linkage: not all verified biological relationships have overlapping demographic footprints in healthcare data. Unlike rule-based approaches, the framework provides probability scores that enable configurable deployment thresholds.

**Conclusion:** The Random Forest models demonstrate high performance suitable for large-scale research deployment and are ready for testing in clinical applications rather than immediate widespread clinical use. The methodology can be adapted to other EHR systems, supporting family-centered study design in patient-centered outcomes research.

## Introduction

Family health history is among the strongest known risk factors for many common diseases, including cardiovascular disease, diabetes, and various cancers.^1^ Understanding patterns of disease within families requires accurate identification of familial relationships within healthcare datasets. Electronic health records (EHRs) contain demographic information that, when properly linked, can reveal family structures and enable research into hereditary disease patterns, shared environmental exposures, and family-level health outcomes. In addition to genetic predisposition shared within families, shared environmental and social factors may support improved prediction of illnesses arising from contagion, toxic exposure, or violence.

The ability to identify family relationships from EHR data has become increasingly important for patient-centered outcomes research (PCOR), epidemiological studies, and clinical care coordination.^2^ Researchers studying hereditary conditions need to identify affected family members to understand transmission, penetrance, and expressivity patterns. Healthcare systems seeking to implement family-centered care coordination require accurate family linkage to identify households with multiple members receiving care.

Current clinical practice relies primarily on patient-reported family history, which has well-documented limitations.^3,4^ Patients often have incomplete knowledge of their family medical history, particularly for more distant relatives. Time constraints during clinical encounters limit the depth of family history documentation, and patient-reported information may contain errors due to recall bias or misunderstanding of medical conditions.

Traditional record linkage methods, designed primarily for identifying duplicate patient records or combining an individual’s data across more than one data source, are insufficient for detecting complex familial relationships.^5^ Deterministic approaches requiring exact matches on specific fields fail to account for the natural variations in demographic information that occur within families over time, such as name changes after marriage, address changes due to residential moves, and variations in how names are recorded across different healthcare encounters.

Previous approaches to family linkage have demonstrated important limitations. Many studies relied on proprietary identifiers or dataset-specific linkage variables that limit generalizability to other healthcare systems.^6,7^ Validation approaches often involved manual review of small samples without systematic evaluation across diverse demographic groups.^8^ The development of generalizable, validated methods for family relationship identification and classification remains an important gap in health informatics research.

This study addresses the gap by developing a machine learning framework that identifies five distinct family relationship types using only standard demographic fields available in most EHR systems. The framework is developed through an iterative, error-driven refinement process and is validated against a gold standard derived from vital records, with additional assessment through temporal shift testing.

### Background and Significance

Research on family relationship identification from healthcare data has evolved through several approaches. Early methods relied primarily on household identifiers available in administrative datasets. Angier and colleagues used Oregon Health Plan household case identifiers to group household members and applied rule-based approaches using age and sex to identify parent-child relationships.^6^ While effective within specific datasets, these approaches depend on identifiers not available in most EHR systems, limiting generalizability.

Polubriaginof and colleagues advanced the field by using emergency contact information recorded in EHR systems to infer family relationships, identifying 7.4 million relationships across three healthcare systems.^9^ This approach demonstrated the feasibility of using routinely collected demographic data for family linkage, and Krefman and colleagues later reimplemented it in Python as P-RIFTEHR, which adds graph-based conflict checking and inference of additional relationships.^10^ Harron and colleagues applied both deterministic and probabilistic record linkage methods to link maternal and infant records, demonstrating the utility of traditional linkage approaches for specific relationship types.^11^ Mayer and colleagues developed methods specifically for identifying twin relationships using birth dates, last names, and supplemental data such as family size and address information obtained from the Marshfield Clinic electronic medical record system,^12^ and more recently applied natural language processing to obituaries to recover family structures for older generations that are typically missed by EHR-derived pedigrees.^13^ Johnson and colleagues created algorithms for estimating household composition and size from co-residence and age differences in the SAIL Databank, a population-scale EHR repository in Wales.^14^ These studies demonstrated the potential for demographic-based family linkage but were designed for limited relationship types, depended on data elements not uniformly available across systems, or required external data sources.

Recent reviews have highlighted the growing use of matching algorithms to create linked relational networks in EHR and health databases. Campbell and colleagues provided an overview of approaches for linking household members and defining relational networks using routine health data, noting applications in both family-level care and intergenerational epidemiology.^2^ The review also identified the need for more rigorous validation approaches and greater attention to generalizability across different healthcare systems and populations.

Several studies have examined family structure identification in specific geographic regions, including Nordic countries,^15–17^ Taiwan,^18^ Manitoba in Canada,^19^ and Western Australia.^20^ While these studies demonstrated successful family linkage in specific contexts, they often relied on data sources or population characteristics that may not generalize to other settings, particularly the diverse US population and the lack of a national healthcare identifier. Hamm and colleagues provided an international review of multigenerational health research using population-based linked databases, highlighting both the potential and challenges of family linkage across different healthcare systems.^21^

More recent work has continued to advance family linkage methods. Angier and colleagues created a linked cohort of children and parents in a large US national EHR dataset, demonstrating scalability of family linkage approaches.^7^ Weaver and colleagues developed an algorithm to link mothers and infants in US commercial healthcare claims databases for pharmacoepidemiology research.^22^ Huang and colleagues developed E-Pedigrees, which reconstructs family pedigrees from EHR demographic and emergency contact data.^23^

A fundamental challenge in developing generalizable family linkage methods has been the lack of gold-standard datasets containing verified family relationships across diverse populations.^8^ Without such datasets, validation relies on manual review of small samples or comparison to incomplete reference data, limiting confidence in performance estimates. This study addresses this challenge by using birth certificate data as a gold standard for validating family relationship identification methods, though this approach is inherently limited to relationship types documented on birth certificates.

### Comparison to Existing Methods

This approach differs from prior work in several respects. RIFTEHR and its Pythonic revision employ deterministic or rule-based matching, producing binary match/no-match decisions without confidence estimation.^9,10^ A machine learning framework instead provides probability scores that enable configurable deployment thresholds. This capability supports tiered processing in which prediction confidence can be tailored to the intended use, so that high-confidence predictions can be automatically accepted, moderate-confidence predictions flagged for manual review, and low-confidence predictions rejected. In addition, prior methods often relied on proprietary identifiers, emergency contact information, or external sources such as obituaries that are not uniformly available across healthcare systems, whereas the present approach uses standard demographic fields (name, date of birth, address, phone number) available in most EHR systems, a highly pragmatic approach that enhances potential for broad adoption.^13,23^

## Materials and Methods

### Study Design and Data Sources

This study used two large-scale healthcare datasets from Indiana. The Indiana Network for Patient Care (INPC) is a health information exchange that aggregates data from over 100 hospitals, clinics, and healthcare providers across Indiana.^24^ The dataset contains approximately 45 million medical records representing 15 million unique individuals, with demographic information including names, dates of birth, addresses, phone numbers, and Social Security Numbers.

The Indiana Natality dataset served as the gold standard for verified family relationships. Maintained by the Indiana State Department of Health, this dataset contains birth certificate records spanning 1970 to 2024. Each birth certificate includes verified mother-child and father-child relationships, enabling the creation of gold-standard relationship pairs. For sibling relationships, we identified pairs of children sharing the same mother. For twin relationships, we identified births with multiple offspring. Partner relationships were inferred from couples appearing together on the same birth certificate. This study was approved by the Indiana University Institutional Review Board (Protocol #22665) with waivers of informed consent and HIPAA authorization.

### Dataset Provenance and Global Identifier

Records from the Natality dataset were linked to INPC records using a Global Identifier assigned during data curation. The Global Identifier was generated by the Regenstrief Institute record linkage mechanism, which resolves records that refer to the same individual across contributing data sources into a single stable identifier.^25^ Both the INPC and Natality datasets were provided to the research team with the Global Identifier already applied, allowing verified Natality relationships to be projected onto the corresponding INPC records. Because the Global Identifier resolves individuals rather than relationships, it was used only to attach gold-standard labels to record pairs and was not itself provided to the Random Forest classifiers as a feature.

### Analytical Pipeline Overview

The analytical pipeline proceeded through data preprocessing, candidate pair generation, record comparison and feature extraction, gold-standard integration, model development, and validation. Positive training cases were obtained directly from Natality-verified relationships projected onto INPC records, while negative cases were generated from a three-tier blocking strategy applied to INPC records. Within candidate blocks, records were compared to produce a fixed set of 14 comparison features, and five Random Forest classifiers were then trained and validated on held-out data.

### Technical Implementation

All Random Forest models were developed using Python 3.12 with scikit-learn 1.4.2 and NumPy 1.24.^26,27^ Model training required 30-90 minutes per model on a CPU-based system with 256 GB RAM.

### Data Preprocessing

Data preprocessing included standardizing name formats, formatting dates, and handling missing values.^5^ Last names were normalized by converting to uppercase, applying Unicode normalization (NFKD) to handle special characters (e.g., converting accented characters such as é to e and ñ to n), and removing common prefixes (e.g., MC, MAC, DE, VAN, VON) and suffixes (e.g., JR, SR, II, III). Date fields were formatted consistently using ISO 8601. Placeholder values commonly used in the INPC dataset (e.g., “UNKNOWN”, “BABY”, “INFANT”, “VOID” for names; “999-99-9999” for SSN) were identified and treated as missing data.

### Positive Case Generation

Positive training cases (confirmed family relationships) were generated by integrating the Natality dataset with INPC records. Records from the Natality dataset containing verified relationships were matched to INPC records using the Global Identifier. For each confirmed relationship pair, we calculated the same 14 comparison features used for negative cases (described below), ensuring a consistent feature representation across positive and negative training examples. Because both positive and negative pairs are represented by the identical feature vector, no feature is available for one class but absent for the other. This approach ensured that positive training examples represented gold-standard verified relationships independent of any blocking process.

### Negative Case Generation and Blocking Strategy

Negative cases (non-relationships) were drawn from INPC candidate pairs that did not match any relationship in the Natality dataset. The primary computational challenge arose from the combinatorial explosion inherent in pairwise record comparison.^5^ With 15 million individuals, a complete pairwise comparison would require approximately 112.5 trillion comparisons, exceeding practical resource constraints.

To address this challenge, we implemented a three-tier blocking strategy exclusively for generating negative candidate pairs. Blocking groups records that share a blocking key so that only likely related records sharing enough demographic similarity to be plausibly confusable with a true relationship are compared. This serves two purposes: It reduces the comparison space, and it ensures the negative pairs are similar enough to true relationships to be informative training examples. Each blocking key begins with a single-letter prefix (N, A, or D) that identifies how the key was produced from a different combination of demographic fields. The three types of blocking keys were generated in parallel using a union approach:

● **Name-based blocking:** N_Soundex(LastName)_LastName(first 6 chars). For example, “Smith” becomes “N_S530_SMITH”. The N prefix marks the key as name-based.
● **Address-based blocking:** A_ZIP3_Address(first 6 chars). For example, ZIP “98765” and address “123 Main St” become “A_987_123MAI”. The A prefix marks the key as address-based.
● **Demographic blocking:** D_Year(DOB)_Sex_Soundex(LastName). For example, DOB “2000-01-01”, sex “M”, and last name “Smith” become “D_2000_M_S530”. The D prefix marks the key as demographic.

A record pair entered the negative candidate set if it shared a blocking key under any of the three schemes. This union strategy maximized recall by ensuring that pairs sharing any relevant characteristics were considered. Requiring negative pairs to share at least one blocking key also produced meaningful negative examples: Given roughly 1.1 million linkable relationship pairs in the final dataset among approximately 112.5 trillion possible pairs, randomly selected pairs would be trivially easy to classify as non-relationships. By requiring a shared blocking key, we generated challenging “close negatives” that share superficial similarities with true relationships, ensuring the classifier learns discriminative patterns beyond the mere absence of shared characteristics.

Blocking alone did not reduce the comparison space sufficiently, because large blocks (for example, common surnames) still generate a quadratic number of within-block comparisons. We therefore applied two further filtering steps to the blocked pairs before any features were computed. First, within each block, records were sorted by blocking key so that similar records were adjacent, and comparisons were restricted to a fixed-size sliding window of 100 records that advanced with a 50% overlap between successive windows. The overlap ensured that pairs spanning a window boundary were still compared, while bounding the number of comparisons per block.

Second, each pair surviving the sliding window was scored with a lightweight preliminary similarity score, assigning 1 point for each of the following conditions: normalized edit distance of last name < 0.5; normalized edit distance of middle name < 0.5; age difference < 55 years; same city; and same state. Pairs scoring ≥ 2 were retained. This threshold was deliberately lenient, functioning as a recall-preserving filter that discards obviously unrelated pairs while retaining any pair with plausible evidence of a relationship.

The edit-distance conditions used the normalized Levenshtein distance:^28^

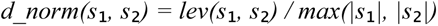

where lev(s₁, s₂) is the Levenshtein edit distance and values are bounded in [0, 1], with 0 indicating identical strings.

Together, these three sequential steps — blocking, sliding-window restriction, and preliminary similarity scoring — reduced the comparison space from over 10¹⁴ possible pairs to approximately 121 million feasible candidate pairs, a reduction of over 99%. Full feature extraction was performed only on the pairs surviving all three steps.

### Feature Extraction

The final feature set comprised 14 variables per record pair, summarized in Table 1. String similarity features included normalized Levenshtein edit distances for last name, middle name, phone number, address, city, and ZIP code. Binary indicators captured Social Security Number agreement, state agreement, and whether the two records differ in sex (sex_difference). The continuous age difference feature measured the absolute difference in years between dates of birth. Sex encoding features (record1_sex, record2_sex) represented biological sex as binary values, and age category features (record1_agecategory, record2_agecategory) classified individuals as Child (< 18 years), Adult (18-50 years), or Older Adult (> 50 years). The sex encoding and age category features were added during Stage 3 (Advanced Feature Engineering) to address systematic misclassification patterns identified in error analysis.

**Table 1.** Complete set of 14 pairwise comparison features used by the Random Forest classifiers. Sex encoding and age category features were introduced during Stage 3 (Advanced Feature Engineering).

| # | Feature | Type | Description |
| --- | --- | --- | --- |
| 1 | edit_dist_lastname | Continuous [0,1] | Normalized Levenshtein distance of last name |
| 2 | edit_dist_middlename | Continuous [0,1] | Normalized Levenshtein distance of middle name |
| 3 | edit_dist_phone | Continuous [0,1] | Normalized Levenshtein distance of phone number |
| 4 | edit_dist_address | Continuous [0,1] | Normalized Levenshtein distance of street address |
| 5 | edit_dist_city | Continuous [0,1] | Normalized Levenshtein distance of city |
| 6 | edit_dist_zip | Continuous [0,1] | Normalized Levenshtein distance of ZIP code |
| 7 | age_difference | Continuous | Absolute difference in years between dates of birth |
| 8 | ssn_match | Binary | 1 if Social Security Numbers agree, else 0 |
| 9 | state_match | Binary | 1 if state of residence agrees, else 0 |
| 10 | record1_sex | Binary | Biological sex of record 1 (Male=0, Female=1) |
| 11 | record2_sex | Binary | Biological sex of record 2 (Male=0, Female=1) |
| 12 | sex_difference | Binary | 1 if the two records differ in sex, else 0 |
| 13 | record1_agecategory | Categorical | Age category of record 1 (Child / Adult / Older Adult) |
| 14 | record2_agecategory | Categorical | Age category of record 2 (Child / Adult / Older Adult) |

### Model Development and Iterative Refinement

We employed Random Forest classifiers for their ability to model non-linear relationships and provide probabilistic predictions.^29^ A Random Forest is an ensemble of B decision trees; each tree is trained on a bootstrap sample of the training data with a random subset of features considered at each split, and the ensemble prediction for a pair x is obtained by averaging the class probabilities of the individual trees:

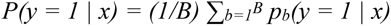

where p_b_(y = 1 | x) is the probability estimate from tree b. The resulting probability was used both to assign the final label at a 0.5 decision threshold and to support the tiered, high-confidence deployment analysis described in the Results. Model development proceeded through four stages, each informed by systematic error analysis of the preceding stage. The stages are summarized in Figure 1 and described below; performance outcomes for each stage are reported in the Results.

**Figure 1.**
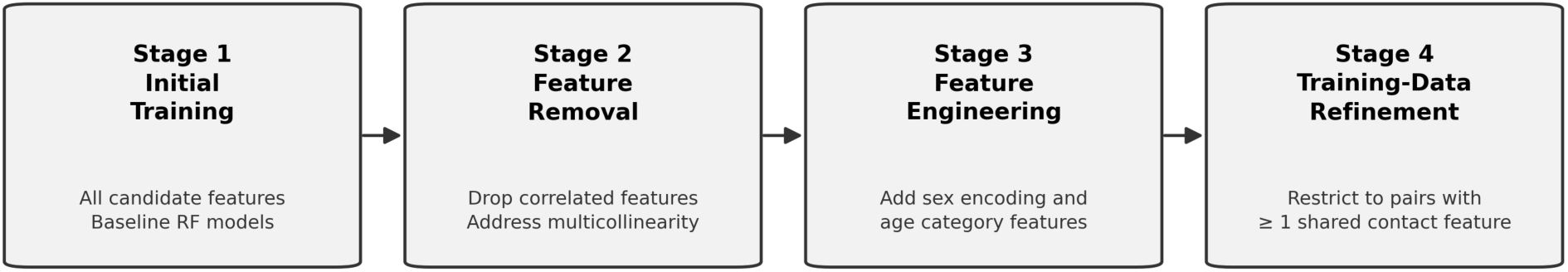
Four-stage iterative model development pipeline. Each stage was informed by systematic error analysis of the preceding stage.

### Stage 1: Initial Model Development

Initial Random Forest models were trained using the complete feature set available at that stage. Analysis of misclassifications on the held-out test set revealed systematic patterns that motivated the subsequent refinement stages.

### Stage 2: Feature Correlation Analysis

Analysis of feature correlations revealed multicollinearity that contributed to model instability. Three features were identified as redundant and removed: edit_dist_dob was highly correlated with age_difference, and the binary matching features (city_match, zip_match) were correlated with their corresponding edit distance features (edit_dist_city, edit_dist_zip). In each case, we retained the more informative feature. For date of birth, age_difference (measured in years) was retained over edit_dist_dob because it directly captures the meaningful biological constraint. For location features, edit distance was retained over binary matching because it provides more granular similarity information than a simple match/no-match indicator.

### Stage 3: Advanced Feature Engineering

Error analysis of the refined models revealed systematic misclassification patterns: False positives in the Mother-Child model were predominantly Father-Child pairs, and vice versa, and the Partner model showed incorrect Partner-Child classifications. To address these patterns, we engineered additional features:

● **Sex encoding features (record1_sex, record2_sex):** Male=0, Female=1, enabling discrimination between maternal and paternal relationships.
● **Age category features (record1_agecategory, record2_agecategory):** Child (< 18), Adult (18-50), Older Adult (> 50), preventing age-improbable cross-generational classifications.

The plausible parent-child age gap is biologically asymmetric — maternal gaps beyond roughly 50 years are effectively impossible, whereas paternal gaps of that size are uncommon but possible — so sex-specific age constraints are a natural refinement for future work. Following feature engineering, hyperparameter optimization was repeated for each model using randomized search with 5-fold stratified cross-validation.^30^

### Stage 4: Training-Data Refinement for Precision Optimization

Analysis of false positives revealed that many misclassifications occurred when the model relied on a single weak feature (e.g., age difference alone) without corroborating evidence. Further investigation of the training data revealed that a substantial proportion of Natality-verified relationships lacked any overlapping demographic features in the INPC records; these pairs had no shared address, phone number, or other contact information that would enable linkage from EHR data alone. We therefore refined the training and test datasets to include only relationship pairs with at least one matching contact feature (A matching contact feature means an exact match (edit distance of zero) on mailing address or on phone number between the two records). This filtering reflects a fundamental constraint: Relationships without any overlapping demographic footprint in the EHR cannot be identified through record linkage, regardless of model sophistication. The refinement reduced the positive class sizes but ensured that the models were trained and evaluated exclusively on linkable relationships. Following this refinement, the models were retrained with the optimized hyperparameters.

### Data Augmentation for Inclusivity

Additional testing revealed false negatives that arose from same-sex partner relationships and gender-field variations in parent records. Because the training data were derived from birth certificates, these cases were systematically underrepresented. To address this bias, we augmented the training data by creating same-sex pair variants from existing opposite-sex pairs, modifying the sex fields of one record to create male-male and female-female versions for approximately 40,000 training records (5% of all training positive cases) in the Mother-Child and Partner models. Augmented pairs were added to the training dataset only, and the held-out test sets contained no augmented data.

### Validation Strategy

Strict separation between training and testing data was maintained through a stratified 70/30 train-test split performed before any model development.^31^ The test set was held separately and used exclusively for final model validation. To confirm that the models learned genuine relationship patterns rather than memorizing specific date ranges, we also conducted temporal shift testing in which all dates of birth were shifted backward by 20 years and model performance was re-evaluated.

## Results

### Test Set Characteristics

All models were evaluated on held-out test sets comprising 30% of the prepared dataset, stratified by relationship type. Table 2 presents the test set composition after training data refinement.

**Table 2.** Test set composition after training data refinement.

| **Model** | **Total Pairs** | **Negative Class** | **Positive Class** |
| --- | --- | --- | --- |
| Mother-Child | 1,465,932 | 1,348,030 (92.0%) | 117,902 (8.0%) |
| Father-Child | 1,420,784 | 1,371,114 (96.5%) | 49,670 (3.5%) |
| Sibling-Sibling | 1,503,177 | 1,391,656 (92.6%) | 111,521 (7.4%) |
| Twin-Twin | 1,446,000 | 1,439,505 (99.6%) | 6,495 (0.4%) |
| Partner-Partner | 1,465,206 | 1,407,907 (96.1%) | 57,299 (3.9%) |

### Model Performance Evolution

Figure 2 presents F1-score trajectories across the four development stages; precision, recall, and F1-score at each stage are given in Table 3. The evolution shows how each refinement stage addressed limitations identified through error analysis. Initial models (Stage 1) produced adequate but suboptimal results, with precision ranging from 0.87 to 0.98 and recall from 0.82 to 0.98. In Stage 2, removal of correlated features paradoxically decreased Mother-Child performance (F1 decreased from 0.86 to 0.81), indicating that additional discriminative features were needed. Stage 3 feature engineering improved performance substantially for the parent-child and partner models, raising F1 score by up to 0.13. Stage 4 (Final Stage) training data refinement improved F1 scores by a further 0.02-0.04 for the Mother-Child, Father-Child, and Sibling models, yielding the final performance metrics reported below.

**Figure 2.**
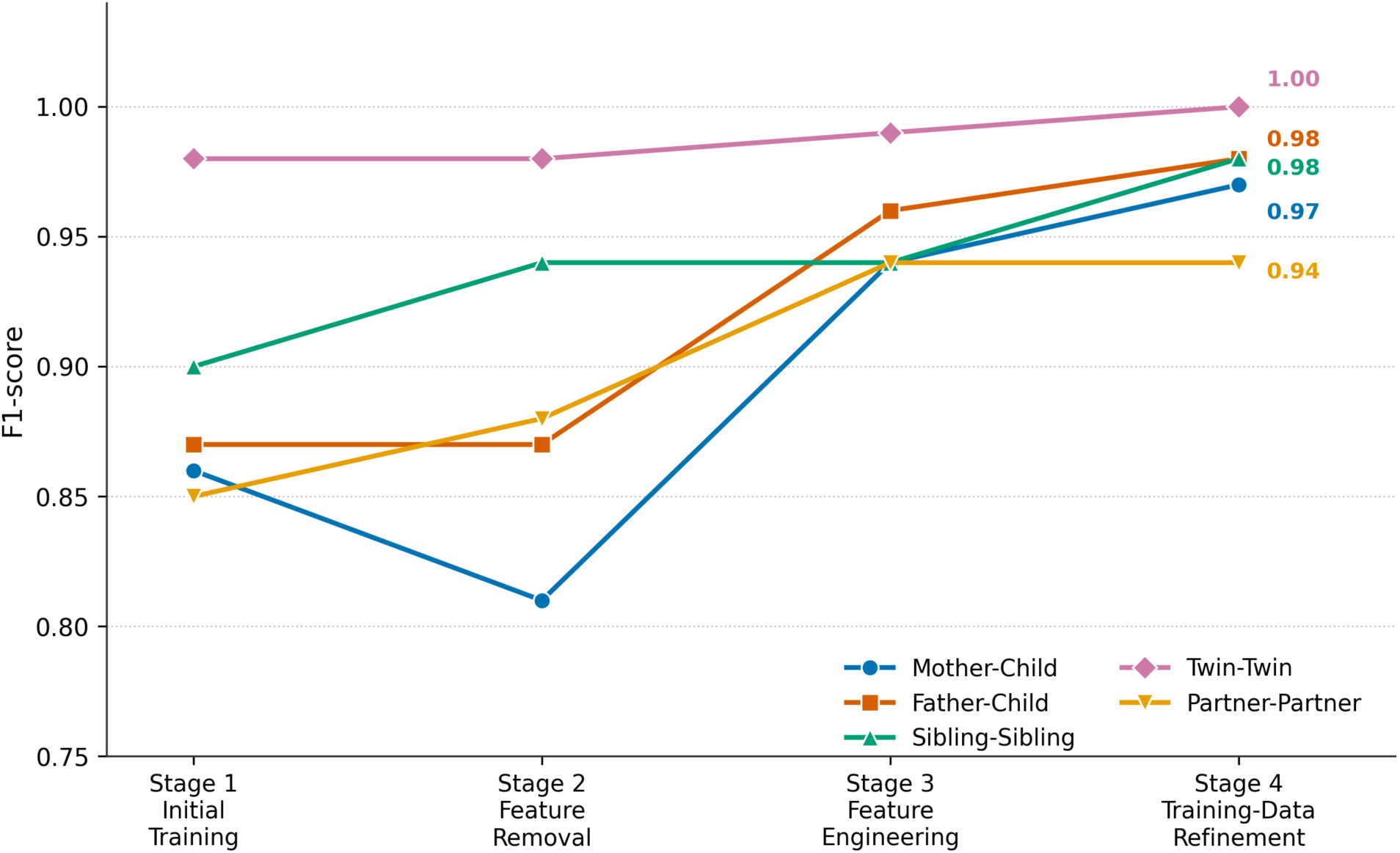
F1-score evolution across four model development stages for five family relationship classifiers.

**Table 3.** Stage-by-stage performance.

| Stage | Mother-Child | Father-Child | Sibling | Twin | Partner |
| --- | --- | --- | --- | --- | --- |
| **Initial Training** | P:0.90<br>R:0.83<br>F1:0.86 | P:0.87<br>R:0.87<br>F1:0.87 | P:0.90<br>R:0.91<br>F1:0.90 | P:0.98<br>R:0.98<br>F1:0.98 | P:0.88<br>R:0.82<br>F1:0.85 |
| **Features Removal** | P:0.85<br>R:0.77<br>F1:0.81 | P:0.84<br>R:0.90<br>F1:0.87 | P:0.93<br>R:0.96<br>F1:0.94 | P:0.98<br>R:0.99<br>F1:0.98 | P:0.90<br>R:0.87<br>F1:0.88 |
| **Feature Engineering** | P:0.94<br>R:0.95<br>F1:0.94 | P:0.95<br>R:0.98<br>F1:0.96 | P:0.94<br>R:0.93<br>F1:0.94 | P:0.99<br>R:0.99<br>F1:0.99 | P:0.93<br>R:0.95<br>F1:0.94 |
| **Training-Data Refinement** | P:0.97<br>R:0.98<br>F1:0.97 | P:0.98<br>R:0.98<br>F1:0.98 | P:0.98<br>R:0.97<br>F1:0.98 | P:1.00<br>R:1.00<br>F1:1.00 | P:0.92<br>R:0.97<br>F1:0.94 |

### Final Model Performance

Table 4 presents the final performance metrics after all four refinement stages (Stage 4 in Figure 2). Models achieved precision of 0.92-1.00, recall of 0.97-1.00, and F1-scores of 0.94-1.00 across all relationship types. The Twin-Twin model achieved the highest performance (F1=1.00), reflecting the near-deterministic signature of twin pairs.

**Table 4.** Final model performance metrics (Stage 4).

| Model | Precision | Recall | F1-Score |
| --- | --- | --- | --- |
| Mother-Child | 0.97 | 0.98 | 0.97 |
| Father-Child | 0.98 | 0.98 | 0.98 |
| Sibling-Sibling | 0.98 | 0.97 | 0.98 |
| Twin-Twin | 1.00 | 1.00 | 1.00 |
| Partner-Partner | 0.92 | 0.97 | 0.94 |

### Confusion Matrix Results

Table 5 presents the confusion matrix results for all models after training-data refinement (*Stage 4)*. The results demonstrate the models’ ability to correctly identify family relationships while maintaining low false positive rates. Per relationship type F1-scores are reported in Table 4.

**Table 5.** Confusion matrix results (true negatives, false positives, false negatives, and true positives) after training-data refinement.

| Model | True Negatives | False Positives | False Negatives | True Positives |
| --- | --- | --- | --- | --- |
| Mother-Child | 1,344,175 | 3,855 | 2,130 | 115,772 |
| Father-Child | 1,370,328 | 786 | 858 | 48,812 |
| Sibling-Sibling | 1,389,774 | 1,882 | 3,447 | 108,074 |
| Twin-Twin | 1,439,491 | 14 | 4 | 6,491 |
| Partner-Partner | 1,402,922 | 4,985 | 1,858 | 55,441 |

### Optimized Hyperparameters

Table 6 presents the optimized hyperparameters for each model, obtained through randomized search with 5-fold stratified cross-validation. Parent-child models required higher max_features values (9-12) than sibling models (4), reflecting the greater complexity of distinguishing parent-child relationships from other adult-child associations.

**Table 6.** Optimized Random Forest hyperparameters for each relationship model. (max_features = number of features considered at each split; min_samples_leaf =minimum samples required at a leaf node; n_estimators = number of trees in the forest)

| Model | max_features | min_samples_leaf | n_estimators |
| --- | --- | --- | --- |
| Mother-Child | 12 | 4 | 600 |
| Father-Child | 9 | 6 | 700 |
| Sibling-Sibling | 4 | 2 | 500 |
| Twin-Twin | 4 | 5 | 500 |
| Partner-Partner | 6 | 5 | 500 |

### Training-Data Refinement Analysis

Across relationship types, 15-78% of Natality-verified relationships lacked any shared contact information in the INPC records and were therefore excluded during training data refinement. This filtering restricted training and evaluation to relationships that can realistically be identified from EHR demographic data.

### Feature Importance Analysis

Figure 3 illustrates feature importance across all five models. Age difference emerged as the most important feature for parent-child and twin relationships, consistent with the characteristic age structures of these relationships. Phone number edit distance was most important for sibling relationships, reflecting shared household contact information. The Partner model showed more distributed importance, consistent with the varied nature of partner relationships.

**Figure 3.**
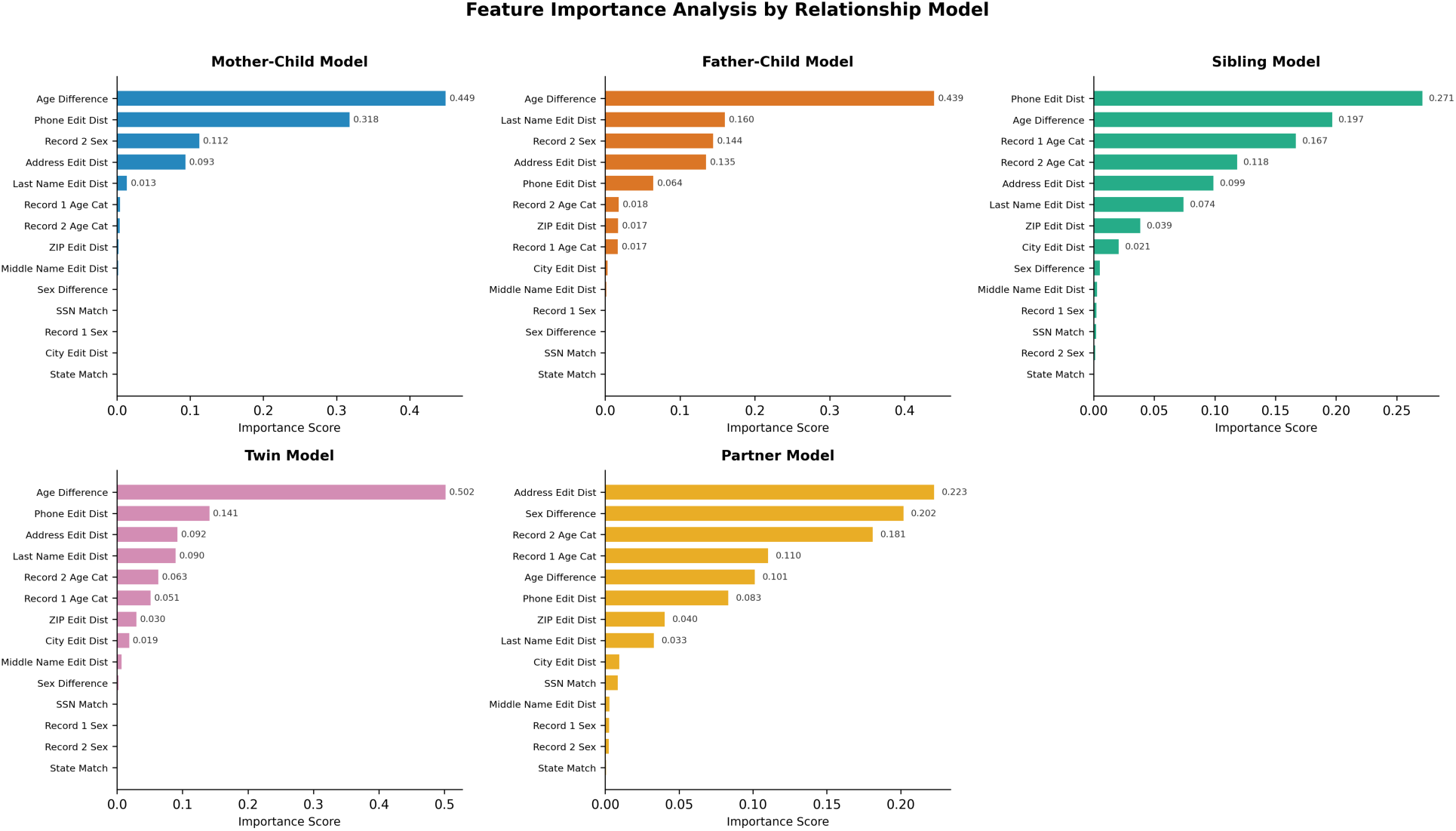
Feature importance analysis for all five relationship models.

### High-Confidence Prediction Analysis

Table 7 presents the distribution of high-confidence true positives (probability ≥ 0.9). These distributions support a 0.9 probability threshold for high-confidence relationship identification, capturing 77-99% of true relationships while minimizing false positives.

**Table 7.** High-confidence true positive distribution (probability ≥ 0.9).

| Model | TP with $P \geq 0.9$ | Percentage |
| --- | --- | --- |
| Mother-Child | 108,569 / 115,772 | 93.8% |
| Father-Child | 47,467 / 48,812 | 97.2% |
| Sibling-Sibling | 105,148 / 108,074 | 97.3% |
| Twin-Twin | 6,399 / 6,491 | 98.6% |
| Partner-Partner | 42,904 / 55,441 | 77.4% |

### Temporal Shift Testing

Temporal shift testing was conducted to confirm that the models learned genuine relationship patterns rather than memorizing specific date ranges. When all dates of birth were shifted backward by 20 years, the models maintained 99% precision despite operating outside the training temporal range, indicating that predictions do not depend on the specific calendar range represented in training.

## Discussion

This study demonstrates the feasibility of accurate family relationship identification from EHR data using machine learning methods. The Random Forest models achieved high performance across all five relationship types, and the four-stage iterative refinement process shows how systematic error analysis can guide methodological improvements.

### Training-Data Refinement

The training-data refinement stage addresses a fundamental constraint in developing and validating EHR-based family linkage methods: Verified biological relationships do not guarantee overlapping demographic data values in healthcare data. Depending on relationship type, only 22-85% of Natality-verified relationships had shared contact information in the INPC records. Filtering to linkable relationships reduced positive class sizes but improved precision for four of the five models, ensuring that models are trained and evaluated on relationships that can realistically be identified from EHR data rather than on cases that would be impossible to detect through record linkage. The Twin model showed the lowest exclusion rate based on shared demographic data (15%), consistent with twins typically receiving care on the same dates, and hence having the same time points for demographic updates and also sharing a household during childhood, while partner pairs showed the highest exclusion rate (78%), consistent with partners receiving care on separate dates, and hence with different time points for demographic updates, and frequently maintaining separate contact information.

### Iterative Refinement and Error Analysis

The four-stage refinement process illustrates how error analysis can guide methodological improvements. Initial models revealed adequate but suboptimal performance. Feature correlation analysis identified multicollinearity, but removal of correlated features unexpectedly decreased Mother-Child performance, motivating feature engineering to add discriminative features. Error analysis showed that false positives in the Mother-Child model were predominantly Father-Child pairs, indicating that additional features were needed to distinguish maternal from paternal relationships; the addition of sex encoding features directly addressed this limitation, and age category features prevented inappropriate cross-generational classifications. The four-stage refinement process raises a natural question about overfitting. Several aspects of the design address this concern. The changes made at each stage were structural rather than data-specific: Removing collinear features, adding sex and age-category features that encode general biological constraints, and applying an a priori linkability criterion based on shared contact information. Temporal shift testing, which displaced all birth dates by 20 years, left precision essentially unchanged, providing evidence that the models learned relationship patterns rather than memorizing the training period. External validation in other healthcare systems would further strengthen confidence in these results.

### Comparison to Existing Methods

These results are consistent with the advantages of a machine learning approach over deterministic methods. The probability scores enabled a tiered deployment strategy, a capability not available in rule-based systems such as RIFTEHR.^9^ The models achieved F1-scores of 0.94-1.00 using only standard demographic fields, without requiring proprietary identifiers or emergency contact information.^6,7^ This capability distinguishes the present framework from prior approaches that produce binary decisions without confidence estimation and that often depend on data elements not uniformly available across healthcare systems.

### Feature Importance Insights

Feature importance patterns align with biological and social expectations for the different relationship types. Age difference was the dominant predictor for parent-child and twin relationships, reflecting their characteristic age structures. For sibling relationships, phone number edit distance was most important, likely reflecting shared household contact information. The Partner model showed more distributed importance, consistent with the varied nature of partner relationships.

### Clinical Deployment Considerations

The high-confidence prediction distributions support a tiered deployment strategy: High confidence (probability ≥ 0.9) for automated acceptance, capturing 77-99% of true relationships; moderate confidence (0.5-0.9) flagged for manual review; and low confidence (< 0.5) for automated rejection. This approach balances efficiency with accuracy, enabling scalable family linkage while maintaining quality. Random Forest feature importance scores also provide model-level interpretability for clinical deployment; because the probability scores were not formally calibrated, deployment thresholds should be re-computed on local data before use; individual predictions can be accompanied by the comparison features that agreed for that pair (for example, “linked as Mother-Child based on a 28-year age difference, shared address, and matching phone number”).^29^

### Applications

This capability extends traditional record linkage beyond individual-level matching to enable population-level studies of hereditary disease transmission, multi-generational health outcome tracking, and family-centered study design in patient-centered outcomes research and epidemiology.^2^ Potential applications include identifying families at high genetic risk for screening programs (with appropriate consent), studying the heritability of complex diseases, identifying family-level risk factors for diseases with a strong contagious or environmental risk, evaluating the impact of family structure and a family-member’s illness on health outcomes for all family members, and designing care coordination to support families and not just individuals.

For PCOR specifically, family linkage affects study design at the point of the data request and the analytic plan. Family structure can be specified as a cohort selection criterion, allowing investigators to request family units rather than unrelated individuals; it supports analyses that treat the family as the unit of observation, including models that account for within-family correlation; and it enables outcome ascertainment across family members, such as evaluating how one member’s diagnosis affects utilization or outcomes for the rest of the household. These design choices need to be specified in advance, because they change what data must be requested and how the analytic dataset is structured.

These applications carry governance implications. The method requires demographic fields — name, address, phone number, date of birth — that are typically excluded from a limited dataset and are not available under a standard de-identified data request. In practice, deployment would require either IRB approval with a waiver of informed consent, or an honest-broker arrangement in which linkage is performed within the covered entity and only de-identified family identifiers are released to investigators. The appropriate model will depend on institutional policy and the intended use. Ethical use of family linkage — including how inferred relationships are disclosed, and the risk of revealing unexpected or unwanted family information — warrants dedicated attention, and we have further work forthcoming on this issue.

### Limitations

This study has several limitations. First, the gold-standard relationships were derived from Indiana birth certificates, which capture only relationship types documented on birth certificates and exclude step-relationships and other household relationships such as roommates. Sibling pairs were defined as children sharing the same mother. Paternal half-siblings — children who share a father but not a mother — are systematically absent from the gold standard, and the sibling model recall is estimated only over maternally linked siblings. The Natality dataset is also incomplete with respect to the true relationships present in the INPC data: Relationships involving individuals whose births were not recorded in Indiana (for example, individuals born in other states or countries), births occurring before the Natality coverage period, and relationships not documented on birth certificates are absent from the gold standard. Consequently, some predictions counted as false positives may represent true relationships not captured in the Natality data, which would cause the reported precision to underestimate the true precision. In addition, preprocessing removed generational suffixes (for example, JR, SR, II, III) during name normalization so that father-son pairs sharing the same base surname would receive an edit distance of zero rather than being penalized for the suffix difference. However, this also removes the one name-based signal that distinguishes a father from a son, and retaining suffix agreement as a separate feature may improve father-son discrimination in future work.

Second, the training-data refinement step restricted training and evaluation to relationship pairs with at least one shared contact feature. This design decision prioritizes precision and reflects real-world linkage constraints, but it means that the reported recall is computed only over linkable relationships. The recall denominator therefore excludes verified relationships that lack any overlapping demographic footprint in the EHR, and the reported recall should not be interpreted as recall over all biologically true relationships. In deployment against unfiltered EHR data, relationships lacking any shared contact feature would be returned as false negatives, and effective recall would be correspondingly lower than the values reported here. This limitation is most relevant when family members move and acquire new addresses, such as after partner separation or when children move out of a parent’s home; our methods also used only the most recent address and addresses in EHRs are updated only when a registration or check-in-type process occurs. The proportion of relationships that fall into this “unlinkable” category warrants further investigation, including the use of historical addresses and address timestamps.

Finally, the models were trained and validated on data from a single state, and validation in other geographic regions and populations would strengthen generalizability claims.

## Conclusion

We developed and validated a machine learning framework for automated family relationship identification from EHR data. The Random Forest models achieved precision of 0.92-1.00 and recall of 0.97-1.00 across the five relationship types after training data refinement. The four-stage iterative refinement methodology shows how systematic error analysis can guide feature engineering that addresses specific misclassification patterns.

Training-data refinement addresses a fundamental constraint in EHR-based family linkage: Not all verified biological relationships have overlapping demographic footprints in healthcare data. By restricting training and evaluation to linkable relationships, four of the five models achieved higher precision while maintaining high recall over those relationships. The framework provides probability scores that enable configurable deployment thresholds, supporting tiered processing for scalable family linkage. The methodology can be adapted to other EHR systems, supporting broader adoption of family linkage methods in health informatics research and enabling new avenues for studying familial health patterns.

## Acknowledgments

We thank the Indiana Health Information Exchange for their support in providing the data that made this study possible. This work was supported by the Patient-Centered Outcomes Research Institute (PCORI). The authors thank Claude (Anthropic) for digital proofreading and grammar polishing during the preparation of this manuscript.

## Author Contributions

Toan C. Ong and Lisa M. Schilling served as principal investigators and provided overall project direction and supervision of the entirety of the work. Abhinav Pundir and Andrew Hill designed, developed, implemented, and evaluated the machine learning models. Michael G. Kahn, Sarah Kautz, Daniel Lindberg, Bethany Kwan, Shaun J. Grannis, and Titus K. Schleyer reviewed the methodology and provided critical feedback on the analytical approach and potential applications and ethical considerations around the use of these methods in PCOR. Shaun J. Grannis and Titus K. Schleyer led the effort to provide patient data from Indiana and facilitated data access through the Indiana Health Information Exchange. Abhinav Pundir drafted the initial manuscript. All authors reviewed, edited, and approved the final manuscript.

## Data Availability

The data underlying this article are sensitive personally identifiable information (PII) and therefore cannot be shared publicly.

